# Heat Stress and PPE during COVID-19: Impact on health care workers’ performance, safety and well-being in NHS settings

**DOI:** 10.1101/2020.09.22.20198820

**Authors:** Sarah L Davey, Ben J Lee, Timothy Robbins, Harpal Randeva, C. Doug Thake

**Author notes:** **Corresponding author**: Sarah Davey.

## Abstract

**Background:** The impermeable nature of PPE worn by health care workers (HCWs) during the SARS-CoV-2 (COVID-19) pandemic can potentiate heat stress which may negatively impact the performance, safety and well-being of HCWs.

**Aim:** The aim of this study was to evaluate perceived levels of heat stress and its consequences in HCWs required to wear PPE during the COVID-19 pandemic in the UK.

**Method:** An anonymous online survey was distributed to HCWs required to wear Type 1 or 2 PPE in NHS settings to evaluate the perceived impact of PPE on: (1) physical and cognitive performance; (2) heat stress and heat-related symptoms; (3) frequency of removing PPE due to discomfort caused by heat stress; and (4) general working-life and well-being.

**Results:** The survey received 224 responses from 192 (85.7%) women and 32 (14.3%) men. Even though 71.9% of respondents wore the less thermally challenging PPE (i.e. Type 2), a median of 3 (IQR: 2,5) heat-related symptoms were reported including syncope (7.7%). A median of 1 (IQR: 0-3) cognitive task was adversely affected with attentional focus being the most affected. To relieve discomfort, 32.6% reported removing PPE on five or more occasions in a shift. Ninety two percent reported PPE made their job more difficult and 76.2% advised that physical performance was impaired. Respondents also highlighted concerns of dermatitis and pressure sores in the facial region (22.3%) amongst other factors.

**Conclusion:** Heat stress experienced when PPE is worn negatively impacts the performance, safety and well-being of HCWs and patients. Therefore, modification to current working practices and current design of PPE is urgently required to improve HCWs’ resilience to pandemics of infectious diseases. Results suggest modifications to the design of the protective face mask and strict enforcement of specific work/rest ratios to limit the duration of PPE use would be immediate impactful interventions.

## 1. Introduction

Since 1^st^ January to mid-August, 2020, there has been a total of 313,798 cases of SARS-CoV-2 virus (i.e. COVID-19) and 41,347 deaths in the UK^1^ presenting a serious challenge to the National Health Service (NHS) who may be considered already under strain. To reduce the risk of health care workers (HCWs) within the NHS workforce from contracting or transmitting COVID-19, HCWs are required to wear Type 1 or Type 2 personal protective equipment (PPE). The extent of protection required is dependent upon HCWs level of contact with patients, the risk stratification of patients for COVID-19, and the type of medical procedure being undertaken (e.g. aerosol generating versus non aerosol generating). Public Health England guidelines state that when carrying out aerosol generating procedures and/or treating high risk patients, Type 1 PPE should be worn i.e. filtering face piece class 3 (FFP3) respirator or surgical mask with integrated visor, disposable fluid repellent coveralls or long-sleeved gowns, full face shield or visor, and disposable gloves. However, when giving direct care, Type 2 PPE is recommended i.e. fluid-resistant (Type IIR) surgical masks (FRSM) or surgical mask with no integrated visor, full face shield/visor/polycarbonate safety spectacles or equivalent, disposable plastic aprons and gloves^2^.

The importance of protecting HCWs and patients from contracting COVID-19 is paramount. However, the impermeable, encapsulating nature of some PPE impedes heat loss, which, when combined with the extra weight of PPE and restricted mobility, can increase the level of heat stress, and as a consequence thermal strain (i.e. raised skin and core temperatures) in HCWs^3,4^, even in cool environments^5,6^.

Heat stress increases the risk of heat-related disorders and is often, but not always^7^, associated with impaired mental processing (e.g. problem solving, decision making, remembering, learning) particularly in complex mental tasks^8,9^. Combining these factors with reduced dexterity, impaired visibility and the occurrence of certain skin conditions through wearing PPE^10-14^, jeopardises the safety, performance and well-being of HCWs with potential consequences for patients. Alongside the continued use of PPE, the incidence of heat waves and the continued rise in global temperature further compounds the potential for HCWs to experience heat stress. In addition, the PPE related discomfort experienced by HCWs has caused concern around adherence to both wearing PPE and the appropriate doffing procedures as it endangers infection control^15,16^. Even though these issues were highlighted as an area of concern during the Ebola virus disease (EVD) outbreak^16,17^, the same issues are clearly evident during the current COVID-19 pandemic^12,13^. This is concerning, especially due to the global spread of the current COVID-19 pandemic affecting a larger number of HCWs over a prolonged period of time. Therefore, it is important to understand the impact of PPE on heat stress in HCWs to inform future interventions designed to mitigate the level of heat stress experienced. The aim of this study was to evaluate perceived levels of heat stress and its consequences in HCWs required to wear PPE during the COVID-19 pandemic in NHS settings.

## 2. Method

An anonymous survey was used to capture the perceived level of heat stress experienced by HCWs required to wear PPE during the COVID-19 pandemic in NHS health care settings. The survey was also used to capture how any level of heat stress influenced HCWs working-life on a physical, cognitive and well-being level. The survey also assessed whether any discomfort caused by heat stress experienced by the HCW when PPE was worn posed an infection risk due to the consequent removal of PPE. A mixed methods approach to distributing the survey was chosen due to the challenges of completing the research during the COVID-19 pandemic. The survey was distributed both in paper and online format within the University Hospital Coventry and Warwickshire NHS trust and other NHS settings (via social media; LinkedIn, Twitter) between May and August 2020. Both Type 1 and Type 2 PPE ensembles (described in the introduction) commonly used in NHS settings were assessed.

More specifically, the questionnaire captured the following data: demographics (age, sex, ethnicity); profession (e.g. physician, nurse, physiotherapist); working environment (e.g. ward, accident and emergency, intensive care unit, rehabilitation); working shift pattern (09:00-17:00, 07:00-20:30, 19:00-07:00, day/night shift rotation); hours per day PPE is worn (0-4 hours, 4-8 hours, 8-12 hours, 12+ hours); how often PPE is changed per shift (never, twice, three times, four times, five times, six times, six+ times); and the length of time either the whole PPE ensemble, protective facial mask (PFM), or apron was worn on each occasion within a shift.

Perceptions of the level of heat stress experienced were evaluated by assessing, retrospectively, temperature sensation (TS) and thermal comfort (TC) when PPE was worn and comparing the change in sweating when PPE was worn to when PPE was not worn. These perceptions were assessed by Likert scales (Supplementary S1). The level and consequence of heat stress experienced by HCWs when wearing PPE was also assessed by the number and type of heat-related illness signs and symptoms experienced (i.e. confusion, dark-coloured urine, dizziness, fainting, fatigue, headache, muscle or abdominal cramps, gastrointestinal disturbance [nausea, vomiting diarrhoea], profusely sweating, rapid heartbeat).

The impact of the level of heat stress, or other factors, experienced when wearing PPE on the physical performance of HCWs was evaluated by HCWs’ perception of whether, and to what degree, wearing PPE impaired their physical performance at work and whether PPE made their job easier or more difficult. Both perceptions were assessed using Likert Scales (Supplementary S1). The impact on cognitive performance was assessed by the number and type of cognitive tasks that were perceived to be adversely affected by wearing PPE. The types of cognitive tasks were: making decisions, solving simple problems, solving complex problems, solving simple numerical calculations, solving complex numerical calculations, retrieving information from long-term memory, retrieving information from short-term memory, attentional focus.

The impact of heat stress experienced when wearing PPE on the risk of infection was evaluated by whether HCWs removed PPE within a shift, and how often i.e. once, a few times (2-5 occasions), often (5-10 occasions), regularly (10+ occasions), due to feelings of discomfort or overheating.

The questionnaire also contained a free text option where the respondent could provide additional information of their experiences of wearing PPE and how these experiences may have impacted their working-life. The questionnaire is available as online supporting information, Supplementary S1. Ethical approval was provided by Coventry University Ethics Committee.

### Statistical analysis

Data values are presented as mean ± SD, median ± IQR, range or percentages. Relationships between the categorical measures of type of PPE (Type 1 or Type 2) and length of time PPE is worn (0-4 hours, 5-8 hours, 8 hours plus) and (1) temperature sensation (hot, warm, slightly warm); (2) thermal discomfort (very uncomfortable, uncomfortable, slightly uncomfortable); (3) sweating discomfort (very uncomfortable, uncomfortable, slightly uncomfortable); (4) changes in sweating (largely increased, increased, slightly increased); (5) number of heat stress symptoms (0-2, 3-5, 6-8); (6) number of cognitive tasks impaired (0-2, 3-5, 6-8); (7) impairments to physical performance (No impairment, slightly impaired, impaired, largely impaired); (8) changes in the level of difficulty of role (more difficult, slightly more difficult, no change, slightly easier, easier); and (9) frequency of removing PPE due to discomfort were assessed using the Pearson Chi Square test. Effect size was assessed using Cramer’s *V*. Commonly used interpretations of Cramer’s *V* is to refer to effect sizes as small (0.1-0.3), medium (0.3-0.5) and large (0.5-1.0)^18^. All statistical procedure were performed using the Statistical Package for the Social Sciences 25.0 for Windows (SPSS, Inc., Chicago, IL, USA). The additional comments were analysed independently by two of the authors using thematic analyses (reflexive approach) with concurring themes being presented^19^.

## 3. Results

The survey received 230 responses. Due to 6 of the respondents reporting being only required to wear a protective face mask (PFM), 224 responses from 192 (85.7%) women and 32 (14.3%) men were included in the analysis of the survey responses (demographic information is provided in Table 1). Due to face covering becoming an emerging theme as affecting the respondents working-life, any additional comments of the 6 respondents who only wore a PFM were included in the thematic analysis (n=4). Therefore, a total of 112 additional comments were included in the thematic analysis.

**Table 1.** Demographics of respondents (n = 224)

|  | Frequency | Percentage (%) |
| --- | --- | --- |
| **Age (years)** |  |  |
| 18-29 | 55 | 24.6 |
| 30-39 | 65 | 29.0 |
| 40-49 | 50 | 22.3 |
| 50-59 | 43 | 19.2 |
| 60+ | 11 | 4.9 |
| **Ethnicity** |  |  |
| English/Welsh/Scottish/NI/British | 187 | 83.5 |
| Irish | 3 | 1.3 |
| Other white | 4 | 1.8 |
| Mixed ethnic background | 7 | 3.1 |
| Indian | 7 | 3.1 |
| Pakistani | 1 | 0.4 |
| Chinese | 5 | 2.2 |
| Asian background | 2 | 0.9 |
| African | 6 | 2.7 |
| Caribbean | 1 | 0.4 |
| Other black/African/Caribbean | 1 | 0.4 |
| **Role** |  |  |
| Allied Health Practitioner | 48 | 21.4 |
| Medic | 30 | 13.4 |
| Nurse/Sister | 78 | 34.8 |
| Health Care Assistant | 40 | 17.9 |
| Admin/managerial/research | 16 | 7.1 |
| Other | 12 | 5.4 |
| **Working environment** |  |  |
| Ward | 119 | 53.1 |
| Accident and emergency | 22 | 9.8 |
| Outpatients | 40 | 17.9 |
| Operating Theatre | 16 | 7.1 |
| ICU/HDU | 14 | 6.3 |
| Community | 7 | 3.1 |
| Office | 3 | 1.3 |
| Reception | 3 | 1.3 |
| **Working pattern** |  |  |
| 09:00-17:00 | 96 | 42.9 |
| 19:00-07:00 (rotational) | 7 | 3.1 |
| 07:00-19:00/19:00-07:00 (rotational) | 63 | 28.1 |
| 07:00-20:30 | 37 | 16.5 |
| Other | 20 | 8.9 |

Footnote: \* includes physiotherapists, physiotherapist assistant, occupational therapists. \*\* other services include radiographers, psychiatrists, dieticians (all patient facing roles).

### Type of PPE and usage

Of the respondents 63/224 (28.1%) wore the Type 1 and 161/224 (71.9%) wore the Type 2 PPE ensemble. The respondents wore either ensemble for 0-4 hours (60/224, 26.8%), 4-8 hours (76/224, 33.9%), 8-12 hours (74/224, 33.0%), 12+ hours (14/224, 6.3%) in a shift. Ninety (40.2%) respondents advised they changed the PPE ensemble 6+ times in a shift and 30/224 (12.4%) respondents changed the ensemble less than 3 times within a shift. The average length of time a full PPE ensemble is worn was 181 ± 192 minutes, with the PFM being worn for 478 ± 287 minutes and the apron 91 ± 198 minutes.

### Perceptions of heat stress

Whilst wearing PPE, 162/223 (72.3%) respondents perceived they felt ‘hot’, 52/223 (23.2%) felt ‘warm’, and 9/223 (4.0%) felt ‘slightly warm’. No respondents reported feeling ‘neutral’ to ‘cold’. Due to their perceptions of temperature sensation, 113/224 (50.4%) respondents perceived they felt ‘very uncomfortable’, 88/224 (39.3%) felt ‘uncomfortable’, 20/224 (8.9%) felt ‘slightly uncomfortable’ the remaining three respondents felt ‘comfortable’ or ‘slightly comfortable’. In regard to sweating, 221/224 (98.7%) reported experiencing an increase in sweating with 115/224 (51.3%) reporting their perception of sweating being ‘largely increased’. A median of 3 (IQR: 2,5; range 0-9) heat-related illness symptoms were reported, with headache being the most reported symptom 177/208 (85.1%), followed by fatigue 142/208 (68.3%), profuse sweating 120/208 (57.7%), dizziness 90/208 (43.3%) and dark-coloured urine 76/208 (36.5%). Sixteen respondents (16/208, 7.7%) experienced fainting and 16 (16/224, 7.1%) experienced no symptoms (Table 2).

**Table 2.** Frequency of heat-related illness symptoms (n = 224).

| Heat-related illness symptom | Frequency | Percentage (%)<sup>*</sup> |
| --- | --- | --- |
| Confusion | 17 | 7.6 |
| Dark-coloured urine | 76 | 33.9 |
| Dizziness | 90 | 40.2 |
| Fainting | 16 | 7.1 |
| Fatigue | 142 | 63.4 |
| Headache | 177 | 79.0 |
| Muscle or abdominal cramps | 18 | 8.0 |
| Nausea, vomiting or diarrhea | 43 | 19.2 |
| Profuse sweating | 122 | 54.5 |
| Rapid heartbeat | 49 | 21.9 |
| No symptoms experienced | 16 | 7.1 |

**Footnote:** \* % relates to the total number of respondents as opposed to the number of people who advised they experienced a heat-related illness symptom.

The amount of hours PPE was worn had a significant association with thermal comfort (χ^2^ (4) = 18.237, p = 0.001, Cramers *V* = 0.206), sweating comfort (χ^2^ (4) = 12.393, p = 0.014, Cramers *V* = 0.170) and the number of heat stress symptoms experienced (χ^2^ (4) = 14.171, p = 0.006, Cramers *V* = 0.182). There were no associations with the type of PPE ensemble worn and perceptions of heat stress or heat-related illness symptoms.

> “……………… I feel like my attention is not fully on the patients as I am partly focussed on coping with wearing PPE. Concentration is hard. On days off I have felt completely exhausted and fatigued, **much** more so than usual. In all honesty it has been the worst thing for me in dealing with the whole pandemic. Even once the surge was over and other departments were getting back to some normality I felt that we in ITU had been forgotten about in our full PPE shift after shift even when we were looking after non-COVID patients. Morale was massively impacted because everyone just felt so drained and negative. ……… I seriously worry about if another wave comes and we go back to donning full PPE full-time because I honestly don’t think I could cope.”
>
> Clinical sister 1

### Risk of infection due to heat stress

In a shift 172 (76.8%) respondents reported that they have removed the PPE in order to relieve discomfort or overheating. Seventy three (32.6%) of these respondents reported they removed the ensemble five or more occasions in a shift to relieve discomfort or overheating, with 13.8% removing the PPE on more than 10 occasions. There was a significant weak association with the number of hours PPE was worn in a shift and the frequency the PPE was removed to improve comfort (χ^2^ (6) = 13.460, p <0.05, Cramers’ *V* = 0.201). No associations were found between the frequency PPE was removed and the type of PPE ensemble worn.

### Performance

Respondents 170/223 (76.2%) reported PPE impaired their physical performance at work. A median of 1 (IQR: 0-3, range, 0-8) of the listed cognitive tasks were perceived as being affected when PPE is worn. Sixty five percent (145/223) of respondents reported at least one cognitive task being affected. Attentional focus was reported by the majority of respondents as being affected 134/145 (92.4%) followed by complex problems 60/145 (41.4%) and making decisions 50/145 (34.5%); Table 3. On the whole, 204/223 (91.5%) of respondents reported PPE made their job more difficult with 19/223 (8.4%) reporting ‘no change’ or ‘slightly easier’. There were no associations found between the type of PPE ensemble worn or the length of time the PPE was worn with any of the performance measures.

**Table 3.** Frequency of cognitive tasks adversely affected when wearing PPE (n = 224)

| Cognitive task | Frequency | Percentage (%)<sup>*</sup> |
| --- | --- | --- |
| Making decisions | 50 | 22.3 |
| Solving simple problems | 24 | 10.7 |
| Solving complex problems | 60 | 26.8 |
| Solving simple numerical calculations | 19 | 8.5 |
| Solving complex numerical calculations | 32 | 14.3 |
| Retrieving information from long-term memory | 29 | 12.9 |
| Retrieving information from short-term memory | 45 | 20.1 |
| Attentional focus | 134 | 59.8 |
| Cognition not affected | 79 | 35.3 |

**Footnote:** \* % relates to the total number of respondents as opposed to the number of people who advised they experienced an impairment in a cognitive task.

> “Close up 3D vision is significantly impaired. On few occasions I had to opt out of regular PPE or abandon the procedure.”
>
> Doctor

> “I wear glasses that are constantly steaming up. I also have varifocal glasses and the mask covers the part of my glasses that I read with. My role involves reading very small font from a machine and struggle to see well. My glasses are constantly smeared dirty from the masks too. This causes headaches and fatigue and I am not as efficient at work”
>
> Nursing Sister

### Additional information

Several themes emerged from the individual comments, with two main issues being commented upon to affect respondents’ working-life: heat stress (28.6%) and individual PPE items i.e. the PFM (42.0%) and the visor (22.3%). Several themes emerged on how these two issues affected respondents’ working-life. Further themes related to operating procedures or certain policies regarding PPE use in health care settings (Figure 1).

**Figure 1.**
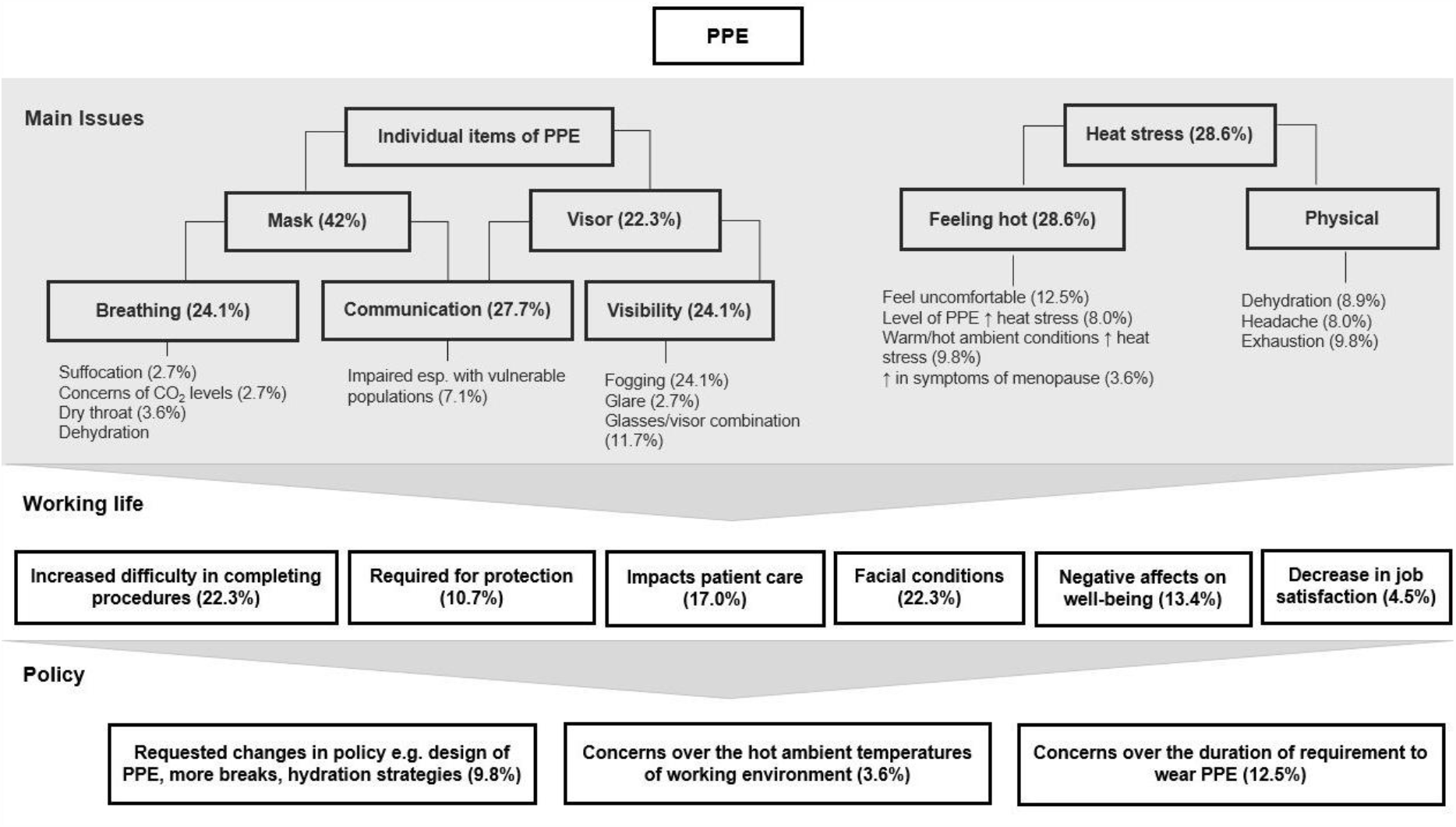
Main themes generated from the additional information provided regarding respondents experiences of wearing PPE and how these experiences may have impacted their working-life (n = 112).

Some respondents who commented on the level of heat stress experienced highlighted that this issue was exasperated by the level of PPE worn (8.0%) and by higher ambient temperatures of their working environment (such as the heatwaves experienced in the UK during the COVID-19 pandemic) (9.8%). Some women (3.6%) also highlighted that wearing PPE increased the severity of symptoms associated with menopause such as hot flushes.

In individuals who commented on issues related to wearing a PFM, 24.1% expressed how the PFM elicited difficulties in breathing, 2.7% reported feelings of suffocation and another 2.7% reported concerns of possible elevated carbon dioxide levels in the mask. Some respondents also reported having breathing difficulties when wearing the visor, but reduced visibility was more of an issue (24.1%). Eye protection steaming up was reported as the main cause for impaired visibility (24.1%). However, a few respondents reported that glare caused by the visors when viewing certain display monitors compromised visibility (2.7%). Several respondents advised that the combination of glasses and wearing goggles/visors/masks was particularly problematic, especially in regard to steaming up (11.7%). Both the mask and visor were also reported to create a barrier that negatively impacted communication with patients and colleagues (27.7%), especially from vulnerable groups such as the elderly, patients diagnosed with a mental disorder or with hearing impairments (7.1%).

> “It is difficult to communicate with those with hearing impairments. It is very difficult to communicate with those with dementia, who would normally read facial expression rather than listening to your speech”
>
> Physiotherapy assistant

Several themes emerged in regard to how the two main issues (i.e. heat stress and individual PPE items) impacted respondents’ working-life. In relation to performance, 22.3% of respondents commented on how PPE increased the difficulty in performing certain procedures with cannulation, CPR, and conducting physiotherapy assessments being specifically mentioned. Personal issues were also reported with some individuals experiencing some kind of contact dermatitis in the facial region (e.g. acne or pressure sores around the nose or ears) (17.9%) or eye irritation (4.5%) (i.e. dry eyes, sore eyes or itchy eyes) due to wearing either the PFM or visor for extended periods of time. Negative effects on individuals’ health and well-being were also reported with 13.4% of respondents advising that PPE affected their physical or mental health including raising levels of stress. A further 4.5% made comments related to experiencing a decrease in job satisfaction due to wearing PPE. However, 10.7% of respondents commented that PPE made them feel safe and protected at work from contracting or transmitting COVID-19 and therefore valued the importance of wearing PPE.

Due to the impact of wearing PPE on some respondents’ working-life, additional themes related to policy emerged. Approximately 10% made comments related to changing policy regarding wearing PPE i.e. scheduling longer breaks, wearing less thermally stressful ensembles, increasing the ability to hydrate, working in cooler environments and not having to wear PPE when treating patients not diagnosed or suspected to have COVID-19.

> “Wearing PPE all day makes me dread attending work. I don’t take work any additional shifts. My communication with patients is severely affected. I feel unwell all shift, can’t concentrate or think straight. I get agitated easily. Unsure why surgical mask required throughout shift.”
>
> Clinical Sister 2

## 4. Discussion

The main findings from the survey were that even though the majority of participants wore the less thermally challenging PPE ensemble (i.e. Type 2), ∼72% of respondents perceived they felt ‘hot’ with ∼99% reporting an increase in sweating (subjective evaluation) and as a consequence ∼98% experiencing some level of discomfort. To relieve these feelings of discomfort or overheating, ∼77% respondents reported that they had removed the PPE in a shift, increasing their risk of infection if not doffed and possibly donned again appropriately. In addition, at least three heat-related illness symptoms had been experienced by respondents whilst wearing PPE, with headache and fatigue being the two most reported symptoms. In regard to performance, ∼76% advised that their physical performance had been impaired when PPE was worn. At least one cognitive task was adversely affected, with attentional focus being the most frequently reported (∼92%). The majority of respondents (∼92%) highlighted that wearing PPE made their job more difficult. However, these difficulties were not solely associated with heat-related illness symptoms, but other ergonomic factors such as the bulkiness of PPE and visual impairment due to goggles or visors steaming up. The PFM and visor were highlighted to be the most problematic items of PPE with the most common issues reported being: (1) difficulties with communication to patients, especially vulnerable patients; (2) impaired visibility; (3) issues in the facial region such as dermatitis (e.g. acne) and pressure sores on the nose and ears; and (4) restricting the ability to adequately hydrate thus increasing the risk of dehydration.

The semipermeable or impermeable nature of some PPE increases insulation and impedes sweat evaporation, limiting the ability to dissipate heat through the body’s normal strategies of heat loss (i.e. sweat evaporation, convection, and radiation). The level of thermal strain experienced when PPE is worn has been well documented in several occupational settings (e.g. Firefighters, Military), including health care settings^6,20,21^. The experiences of HCWs wearing PPE during the EVD outbreak in West Africa 2014-2016 created a resurgence of interest in this topic in health care settings. In their laboratory based study, Coca et al. (3) evaluated the physiological and subjective responses to wearing three different types of PPE typically worn during the 2014-2016 EVD outbreak. The three ensembles (E1, E2 and E3) differed in regard to thermal resistance, but E1 and E2 could be considered similar to the Type 1 PPE ensemble evaluated in the present study whilst acknowledging that thermal properties of clothing can vary widely between suppliers and designs. During 60 minutes of light exercise (i.e. continuous walking, 2.5mph, 0% grade) in environmental conditions of 32 °C, 92% relative humidity (rh), moderate to high levels of thermal strain was experienced with the magnitude of strain being influenced by the level of thermal resistance provided by the PPE (i.e. core temperature: 38.20, 38.80, 38.90 °C, heart rate:136, 156, 163 beats.min^-1^, % of body mass lost due to sweat secretion: 1.3, 1.7, 2%, in E1, E2 and E3, respectively).

In field studies, the level of thermal strain measured in HCWs wearing PPE during the EVD outbreak was less severe^4,21^. For example, over a similar average exposure time i.e. ∼65-75 minutes, the group mean core temperature was raised to 38.0 °C with 20-50% of individuals experiencing core temperatures above 38.5 °C. The lower level of thermal strain could be explained by the less thermally stressful environmental conditions experienced in these field studies (i.e. ∼ 29-30 °C, 50-65% rh) in addition to the HCWs involved in the field studies being: 1) acclimated to the environmental conditions; 2) able to self-pace their physical activity; and 3) performing intermittent vs. continuous physical activity. Regardless, core temperatures measured in these studies have been associated with both physical and cognitive impairments and mild symptoms of heat-related illnesses i.e. fatigue, headache, nausea, dizziness^20,22^. The environmental conditions of these studies are similar to those that have been recorded in some healthcare settings in the UK (e.g. 28-33 °C), especially in old hospital buildings that do not have air conditioning^23,24^ and are typical of the ambient temperatures reached during the heatwaves experienced in the UK between May and August, 2020. Therefore, these levels of thermal strain are likely to have been experienced by HCWs during the COVID-19 pandemic in just one hour of their shift and may partly explain the prevalence of heat-related illness symptoms and impairments in cognition reported in the present study.

Other survey based studies conducted either during the 2014-2016 EVD pandemic^11^ or during COVID-19^14,25,26^ concur with the results of the present study, with heat and dehydration being highlighted as significant issues for HCWs when PPE is worn. Interestingly, in one study that evaluated the prevalence and severity of adverse reactions related to heat stress in front-line healthcare professionals (doctors and nurses) to PPE, the proportion of respondents experiencing heat stress was slightly less than that found in the present study (i.e. 92.9% vs. 68.2%)^14^. However, this may be due to the HCWs in the present study wearing similar PPE ensembles for a longer period of time with ∼73% of respondents wearing PPE for longer than four hours in a shift. In addition, the present study included other roles such as health care assistants who reported more time spent wearing PPE than medics. Medics may have more control over their working pattern compared to other roles and therefore more able to abide to certain policies designed to mitigate heat stress when PPE is worn (e.g. specified work/rest ratios and maintaining hydrated).

In the present study, in regard to cognitive performance, approximately two thirds of the respondents (∼65%) reported one or more cognitive tasks being impaired by wearing PPE. Of the cognitive tasks assessed, the complex cognitive tasks were more often reported to be affected i.e. attentional focus (95.7%), solving complex problems (32.6%), and making decisions (28.3%). These findings are not surprising as it is widely acknowledged that raised core and/or skin temperatures can impair cognition and that simple cognitive tasks are less vulnerable to heat stress than more complex cognitive tasks^8,22,27,28^.

The perception of thermal discomfort has also been associated with impaired cognition^27^. In the present study, ∼81% of respondents experienced some level of discomfort when PPE was worn which may partly explain the impact PPE had on the cognitive tasks assessed. Regardless of the cause, this impairment in cognition may not only affect performance, but also compromises the health and safety of HCWs and patients. Not only has the prevalence of occupational heat stress been strongly associated with workplace accident rates^29^, the majority of accidents reported in certain workplaces has also been found to occur in the hotter summer months^30^. In addition, discomfort has been identified as a factor that may compromise infection control due to frequent donning and doffing of PPE that maybe completed incorrectly^16,31^. In the present study, ∼77% respondents stated they removed the PPE due to discomfort or feelings of overheating. Unfortunately, we cannot ascertain whether this frequent donning and doffing increased the transmission of COVID-19 amongst HCWs. However, donning and doffing can take time if completed correctly^31^, which will negatively affect the productivity of HCWs; a factor highlighted in the additional comments.

In the present study, ∼76% of respondents reported PPE impaired their physical performance at work and ∼92% of respondents reported PPE made their job more difficult. Previous studies have highlighted that completing medical tasks can be adversely affected when PPE is worn with the length of time taken to complete the task more often being affected rather than the successful completion of the task^32-35^. This is supported by some of the respondents highlighting that certain tasks such as cannulation being made more difficult rather than not being able to complete the task. The increase in time taken to complete a task generally occurs as individuals regulate their pace of work to a lower intensity to reduce heat production and/or due to cognition being impaired by heat stress^36^. Regardless, if tasks take longer to complete, the productivity of HCWs will be compromised.

Several factors have been highlighted to impair or increase the difficulty of certain tasks in health care settings with: (1) reduced visibility caused by eye protection^11,25^; (2) difficulties in effective communication caused by the PFM and/or visor^11,25^; and (3) reduced dexterity caused by the gloves^10^ being highlighted as the most common factors. In the present study the PFM appeared to be the most problematic, with compromising communication being the main complaint, especially when treating patients from vulnerable populations (e.g. elderly, patients with learning difficulties, mental disorders or impaired hearing). Issues with wearing a PFM over long periods of time have been previously highlighted, with PFM use found to cause certain skin conditions, breathing difficulties and increased levels of thermal discomfort^13,14^.

Increases in thermal discomfort when wearing a PFM may be explained by the facial region being recognised as being highly thermosensitive^37,38^. For example, Cotter & Taylor (38) demonstrated that cooling the face is 2-5 times more affective in relieving thermal discomfort than any other equivalent dermal region on the body. Consequently, any increases in the temperature and/or humidity of the PFM’s microclimate may have more of an impact on thermal sensation rather than body temperature per se^38^. Therefore, improvements in the design of the PFM and/or visor to reduce the temperature and/or relative humidity of the PFMs microclimate may assist in reducing both overall discomfort and issues related to visibility (i.e. steaming of eye protection).

As already mentioned, a large proportion of the cohort in the present study (i.e. ∼93%) reported experiencing heat-related illness symptoms. Chronic exposure to heat stress (i.e. over a working day/week) have been associated with medical conditions such as acute kidney disease, especially if chronically dehydrated^39^. ‘Hangover’ like symptoms have also been reported to occur amongst workers who are chronically exposed to thermally stressful conditions, which have the potential to affect individuals’ sleep, appetite and relationships with friends and family^40^. The comments provided by the respondents highlight the impact of wearing PPE on HCWs’ health and well-being with 13.4% reporting that their physical and mental well-being had been affected. Some of these respondents highlighted that they “dreaded going to work” due to the requirement of wearing PPE and some were unsure if they could cope with a 2^nd^ wave in the COVID-19 pandemic if changes aren’t made in regard to policy of PPE use. Furthermore, the additional comments also suggest that certain populations of HCWs may be affected more than others, for example, individuals going through menopause.

The prevalence, level and impact of heat stress experienced by HCWs when PPE is worn is not a new issue and recommendations on how to lower the thermal burden, and associated levels of discomfort, have been provided. These recommendations include modifications to either the design of PPE (e.g. the use of more breathable materials, inclusion of cooling garments) or changes in working practices (e.g. adopting specific work/rest patterns, hydration strategies, pre-cooling the PPE prior to use, pre-cooling individuals before a shift, the use of cooling rooms during breaks and adoption of communication-enhancing technologies^15,41-43^. The present study highlights that this issue still remains, warranting further research in this area to inform modifications to policy and/or current designs of PPE.

### Limitations

There are some limitations to this study. The more thermally challenging PPE ensemble (i.e. Type 1) is underrepresented in the sample population therefore the extent of the heat stress and its impact on HCWs working-life may not be fully represented. Further studies detailing the experiences of HCWs in intensive care environments may provide this information. In addition, it is not understood whether respondents adhered to certain policies designed to alleviate heat stress when wearing PPE which may have influenced individuals’ responses. Males are also underrepresented in the sample population. It is acknowledged that sex-related differences in thermoregulatory responses are present when PPE is worn^44^, therefore, male HCWs’ experiences may differ to that represented in the present study.

## 5. Conclusion

It is generally accepted that PPE is necessary to reduce the transmission of certain infectious diseases and its protective nature will impede heat loss and mobility to some extent. However, the present study highlights that modifications to the current design of PPE, or policies on the use of PPE in health care settings, are urgently required to reduce the level of heat stress and prevalence of other issues that jeopardise the performance, safety and well-being of HCWs and patients. This requirement is reinforced by some of the respondents’ comments which suggest that, due to negative experiences of PPE use during the COVID-19 pandemic, some HCWs may not be resilient to respond to another pandemic or a 2^nd^ wave in the current pandemic. Modifications in the design, or use, of the PFM and visor could be quite impactful in alleviating some of the discomfort and impaired performance experienced by HCWs. Modifying the length of time PPE is required to be worn within a shift or providing cooler working environments could also be implemented to improve the working-life of HCWs when PPE is required.

## Data Availability

The raw data supporting the conclusions of this article will be made available by the authors, without undue reservation, to any qualified researcher.

## Authors’ contributions

The study was designed by SD, TR, C.DT and HR. Data were collected by SD and TR. Data was analysed by SD and BL. Data interpretation and manuscript preparation were undertaken by SD, BL, TR and C.DT. All authors approved the final version of the paper.

## Acknowledgments

The authors would like to thank the participants who volunteered their time to partake in this study.

